# Data-driven prognosis for COVID-19 patients based on symptoms and age

**DOI:** 10.1101/2021.11.08.21266091

**Authors:** Subhendu Paul, Emmanuel Lorin

## Abstract

In this article, we develop an algorithm and a computational code to extract, analyze and compress the relevant information from the publicly available database of Canadian COVID-19 patients. We digitize the symptoms, that is, we assign a label*/*code as an integer variable for all possible combinations of various symptoms. We introduce a digital code for individual patient and divide all patients into a myriad of groups based on symptoms and age. In addition, we develop an electronic application (app) that allows for a rapid digital prognosis of COVID-19 patients, and provides individual patient prognosis on chance of recovery, average recovery period, etc. using the information, extracted from the database. This tool is aimed to assist health specialists in their decision regarding COVID-19 patients, based on symptoms and age of the patient. This novel approach can be used to develop similar applications for other diseases.

## Introduction

The novel coronavirus disease 2019 (COVID-19), the current pandemic, caused by the deadly virus SARS-CoV-2 continues to pose acritical and urgent threats to global health. The outbreak reported first time in early December 2019 in the Hubei province of the People’s Republic of China has spread worldwide^1^. As of 26th October 2021, the overall number of patients confirmed to have the disease has exceeded 245,121,000, in more than 220 countries, though the number of people infected is probably much higher, and more than 4.9 million people have died from COVID-19^2^. This pandemic continues to challenge healthcare systems worldwide in many aspects, including the demands for hospital beds and critical shortages in medical equipment. Thus, the capacity for immediate clinical decisions and effective usage of health-care resources is crucial.

There are numerous analytical and computational studies based on mathematical models, involving Ordinary Differential Equations (ODE)^3–13^ as well as Delay Differential Equations (DDE)^14–21^, to calculate the basic reproduction number *R*_0_ and understand the underlying dynamics of the epidemic. In addition to those mathematical approaches, there are several statistical studies^22–30^, based on various samples of patients such as severe, non-severe, ICU, non-ICU, large size, small size, meta-analysis, estimated the recovery time of the current pandemic. Furthermore, effective screening enables quick and efficient prognosis of COVID-19 and can mitigate the burden on health-care systems. Prediction models that combine several features to estimate the risk of infection have been developed, in the hope of assisting medical staff worldwide in trigging patients, especially in the context of limited health-care resources. These models use features such as computer tomography (CT) scans^31–35^, clinical symptoms^36^, laboratory tests^37,38^, and an integration of these features^39^. Several research papers demonstrate^40^ the general scenario of the COVID-19 symptoms based on age, gender, etc. However, so far, none of them refer to “individual patients”. The goal of the work is to provide a rapid digital prognosis for individual COVID-19 patients.

In the present work, we extract the useful information such as hospitalization, average recovery period, etc. from the database^41^ of Canadian COVID-19 patients. The data file contains the information of 666237 patients, although numerous case histories are incomplete, and we merely consider the entirely reported cases. Here entirely reported cases indicate those cases which provide all the required information such as age group, symptoms, hospitalization status, etc. The database provides eight different age groups; those are age groups 1: 0 to 19 years, age group 2: 20 to 29 years, age group 3: 30 to 39 years, age group 4: 40 to 49 years, age group 5: 50 to 59 years, age group 6: 60 to 69 years, age group 7: 70 to 79 years and age group 8: 80 years or older. We consider eight symptoms; these are: sore throat, headache, runny nose, pain, weakness, cough, fever and shortness of breath. We consider two states of each symptom, either presence or absence, and for those eight symptoms, we obtain a total variation of (2^8^ *-* 1) = 255; neglect the case when none of them are present.

For each variation of those eight symptoms, we associate a label (as an integer), defined as a symptom label. For eight symptoms i.e., 255 combinations along with eight different age groups, we can classify all patients into 8 *×* 255 = 2040 groups. The eight age groups, correspond to the numbers 1, 2, …, 8, and the binary code of the symptoms together can be expressed as a patient digital code. A detailed description about symptom label and patient digital code can be found in the methods section.

From the available database we extract the information for 2040 distinct patient groups, such as hospitalization status, average recovery period, etc. Using those information of 2040 distinct patient groups, we develop a standalone application (app) for rapid digital prognosis for individual patient. The app generates i) the probability for patients of being admitted to the Intensive Care Units (ICU), ii) the plausibility of being admitted to the hospital but not to ICU, iii) the expectation of recovering, and iv) the average recovery periods of the patients. The database provides the scatter diagrams for the desired information as a function of symptom label, so we build a prediction model to obtain the smooth outcomes. We first develop a computer code to extract, to analyze and to compress the relevant information from the database. Secondly, we design the app to include all the required information. The application is available online to anyone.

Due to lack of data for more than 2040 patient groups, we have to confine our study to eight symptoms and without considering the patient gender. For a sufficiently large database of medical records of any disease, we can develop a similar application following the proposed methodology.

## Results

In this section, we describe the outcomes (Figs. 1 to 5), the percentage of admitted to the ICU as well as to the hospital but not to ICU, the percentage of recovered individuals and the average recovery period, obtained from the database^41^, a collection of age, gender, symptoms, hospitalization status (ICU or non ICU), recovery rate, recovery periods, etc. of Canadian COVID-19 patients. Those information help to find the likelihood factors, hospitalization status, recovery period, etc. for new patients.

The black dots (Fig. 1), obtained directly from the database^41^, represent the rate of ICU admission as a function of symptom label, 1, 2, …, 255, for eight different age groups. We also include (in Fig. 1) the calculated results from the prediction model i.e., average, low and high percentages which are represented by the blue, green and red curves, respectively. The positive correlated scatter diagram implies that as symptom label increases, the percentage of admitted to the ICU increases as well. It is also observed that as the age group number increases, the percentage of admitted to the ICU also increases. As an illustration, consider a group of patients with symptom label 249 and ages 60 to 69; the figure of age group 6 provides the ICU related information of this group. According to database^41^, 20% patients in this group were admitted to the ICU. As par prediction model, on average 19.99 % patients in this group were admitted to the ICU, and the range obtained from the prediction model is 8.679 % to 42.23 %.

**Figure 1.**
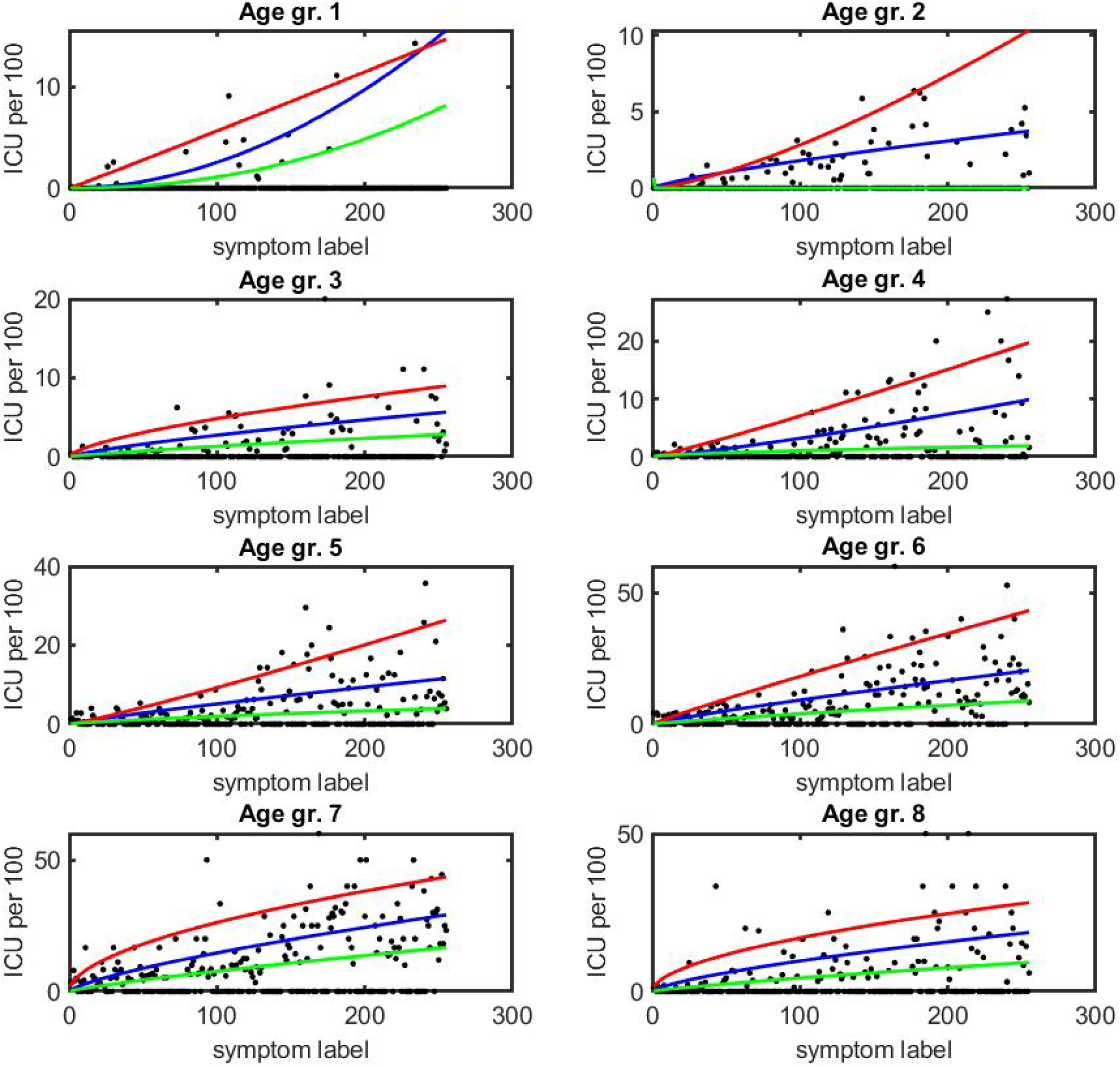
The percentage of being admitted to the ICU as a function of symptom label for different age groups are presented by black dots. The curves blue, green and red indicate the average, low and high probability, obtained from the prediction model.

The black dots (Fig. 2), obtained directly from the database^41^, represent the rate of admission to the hospital but no to ICU as a function of symptom label, 1, 2, …, 255, for eight different age groups. We also include (in Fig. 2) the calculated results from the prediction model i.e., average, low and high percentages which are represented by the blue, green and red curves, respectively. The positive correlated scatter diagram implies that as symptom label increases, the percentage of admitted to the hospital but not to ICU increases as well. It is also observed that as the age group number increases, the percentage of admitted to the hospital but not to ICU increases as well. As an illustration, consider a group of patients with symptom label 249 and ages 60 to 69; the figure of age group 6 provides the hospitalized related information of this group. according to database^41^, 20% patients in this group were admitted to the hospital but not to ICU. As par prediction model, on average 23.95% patients in this group were admitted to the hospital but not to ICU, and the range obtained from the prediction model is 8.244% to 41.29%.

**Figure 2.**
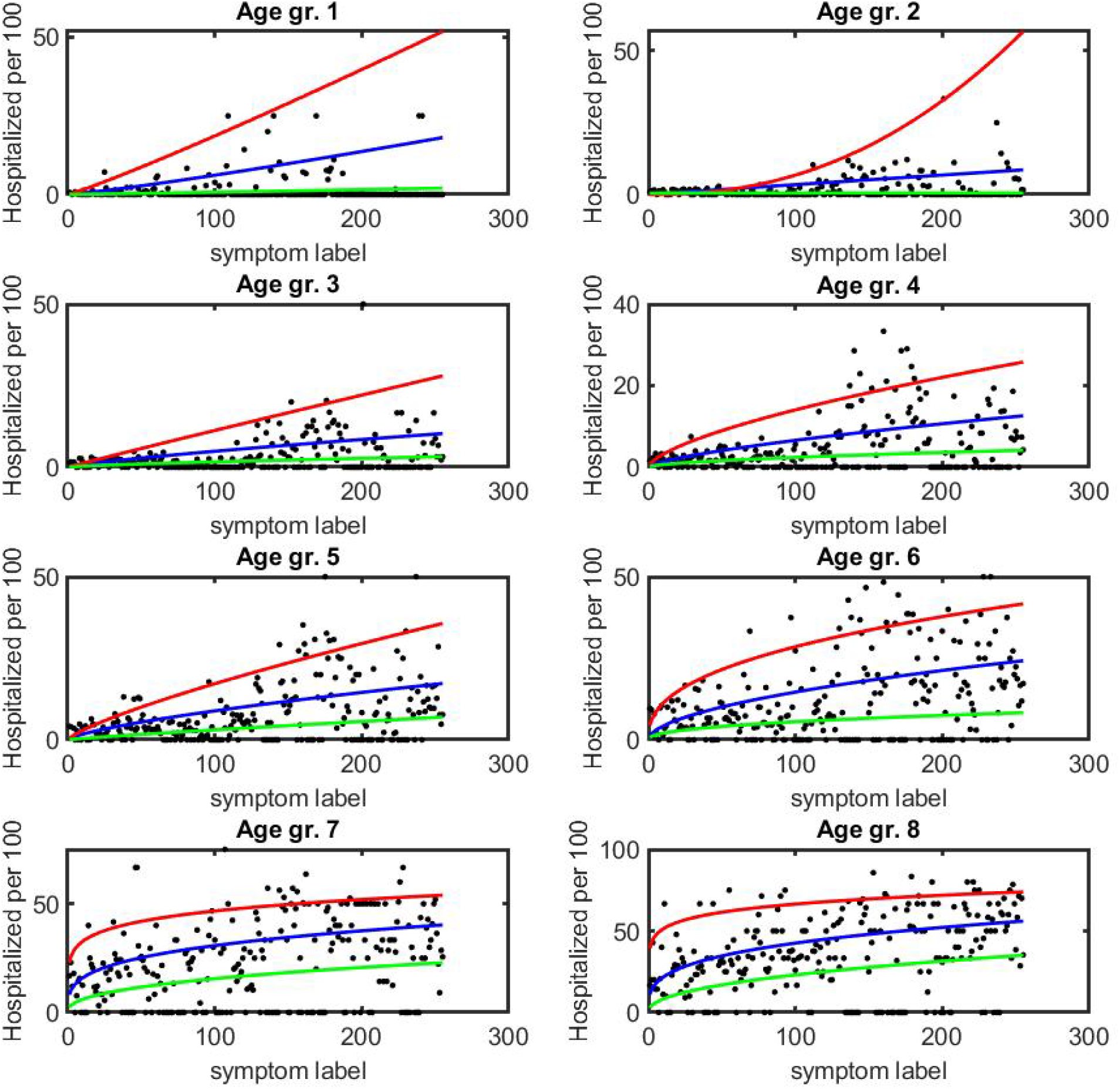
The percentage of being admitted to the hospital but not to ICU as a function of symptom label for different age groups are presented by black dots. The curves blue, green and red indicate the average, low and high probability, obtained from the prediction model.

The black dots (Fig. 3), obtained directly from the database^41^, show the rate of recovered individuals as a function of symptom label, 1, 2, …, 255, for eight different age groups. We observe 100% recovery rate for age groups 1, 2, 3 and 4. The recovery rate has started to decrease from the age group 5, and in some cases for age group 8 the recovery rates are below 50%. We also include (in Fig. 3) the calculated results from the prediction model i.e., average, and low percentages which are represented by the blue and red curves, respectively, for the age groups 5 to 8. The high recovery rate for all age groups with any symptom is 100%. The negative correlated scatter diagram implies that as symptom label increases, the rate of recovery decreases. It is also observed that as the age group number increases, the rate of recovery decreases as well. As an illustration, consider a group of patients with symptom label 249 and ages 60 to 69; the figure of age group 6 provides the information recovery of this group. According to database^41^ 100% patients recovered. As par prediction model, on average 96.56 % patients recovered, and range obtained from the prediction model is 82.16% to 100%.

**Figure 3.**
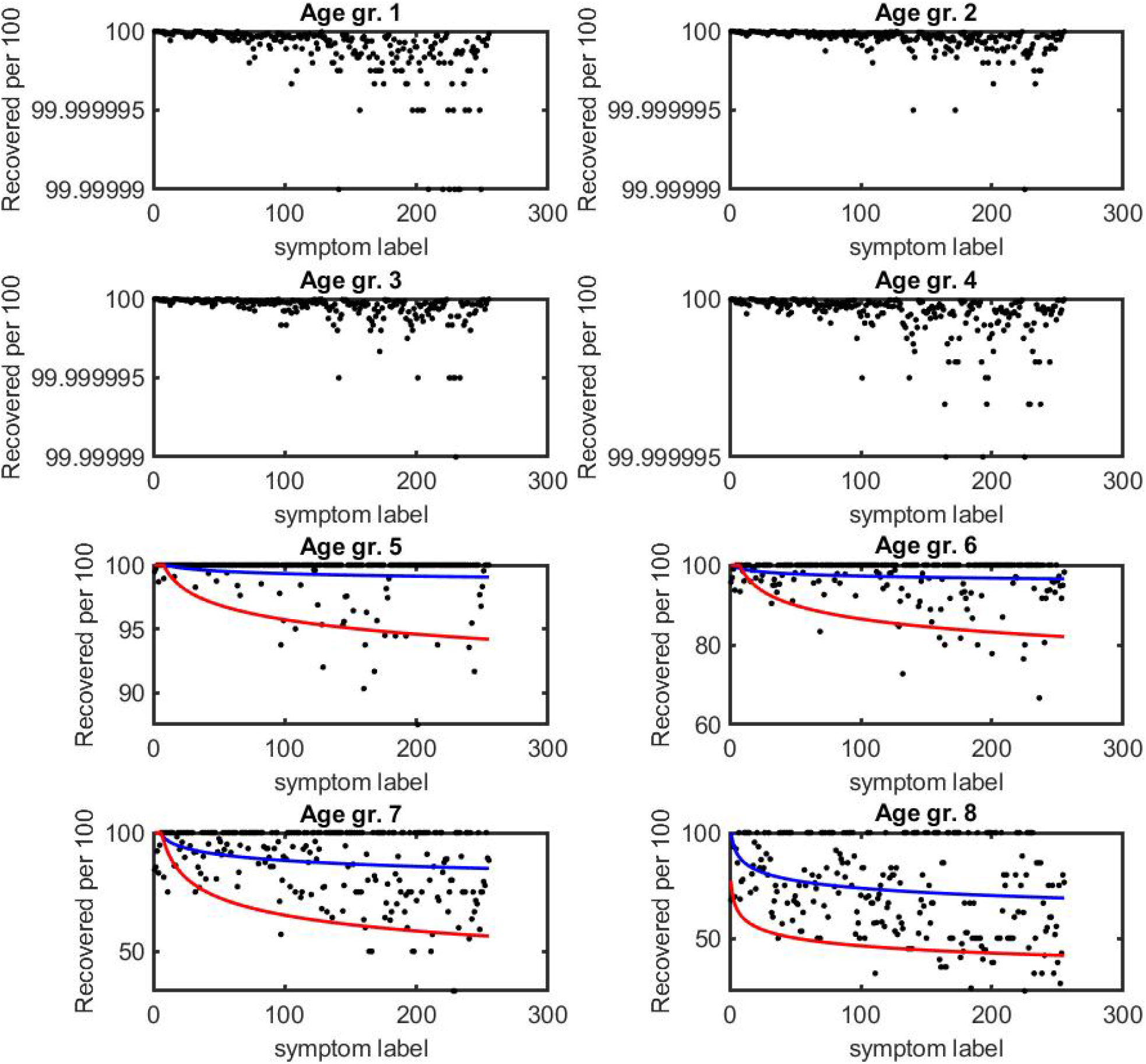
The percentage of recovered individuals as a function of symptom label for different age groups are presented by black dots. The curves blue and red indicate the average and low probabilities, obtained from the prediction model, because the highest probability is 100%.

The black dots (Fig. 4), obtained directly from the database^41^, show the average recovery period as a function of symptom label, 1, 2, …, 255, for eight different age groups. We also include (in Fig. 4) the calculated results from the prediction model i.e., average, low and high recovery periods which are represented by the blue, green and red curves, respectively. The positive correlated scatter diagram implies that as symptom label increases, the duration of average recovery period increases as well. It is also observed that as the age group number increases, the continuation of average recovery period increases as well. As an illustration, consider a group of patients with symptom label 249 and ages 60 to 69; the figure of age group 6 provides the average recovery period of this group. According to database^41^, the patient’s recovery period 2.353 weeks. As par prediction model, the patient’s recovery period on average 2,599 weeks and the range obtained from the prediction model is 1.734 to 3.793 weeks.

**Figure 4.**
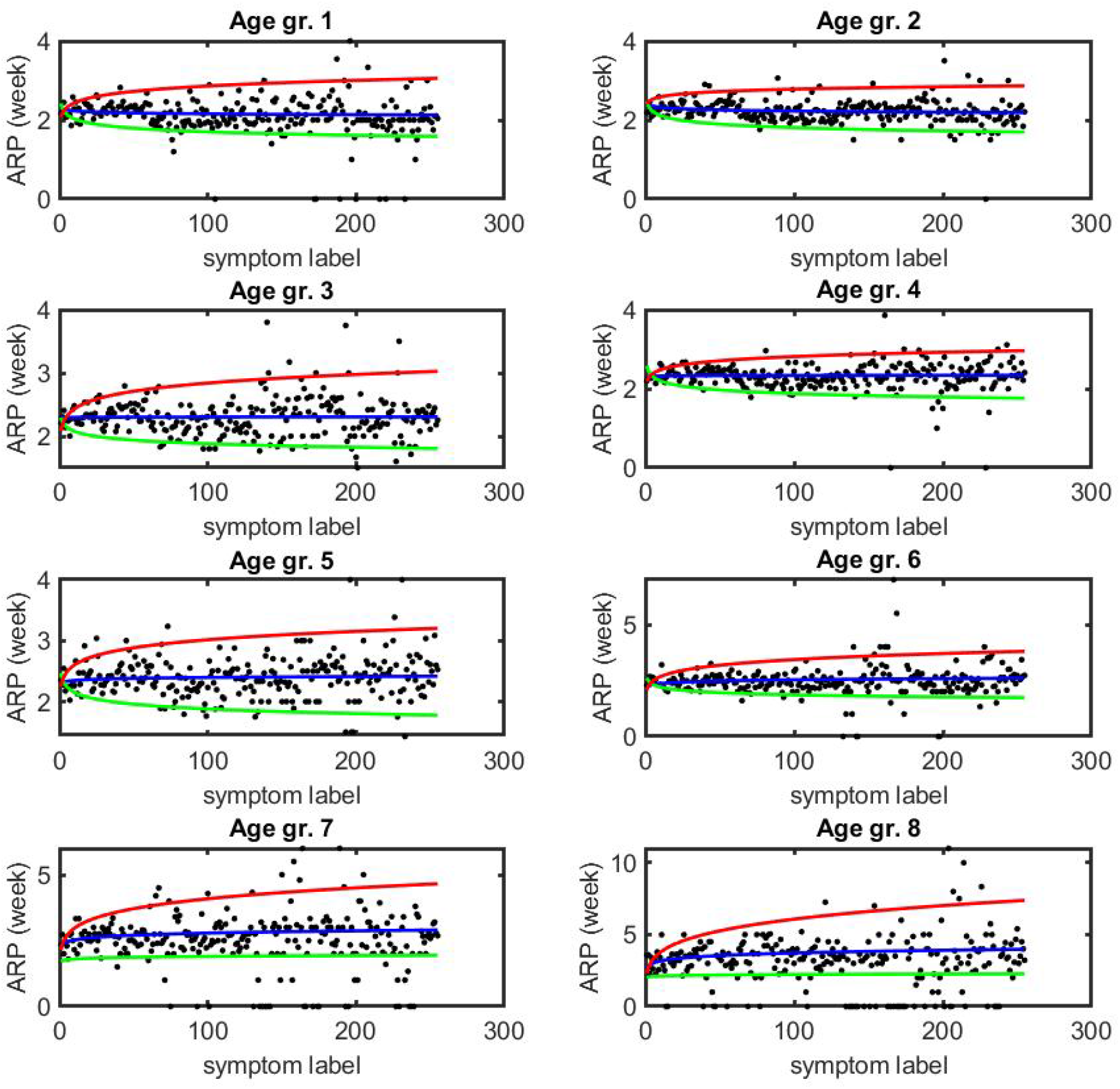
The average recovery periods (in weeks) of the patients as a function of symptom label for different age groups are presented by black dots. The curves blue, green and red indicate the average, low and high probability, obtained from the prediction model.

Employing the information (Figs. 1 to 4) based on Canadian patients, we generate an application for digital prognosis of COVID-19 patient. The sketch of the proposed application (Fig. 5) shows that there are several blocks for input data, age and eight symptoms, and corresponding the output values, several likelihood factors. For example, suppose a 68 years old patient has had the symptoms of sore throat, pain, weakness, cough, fever and shortness of breath. The patient belongs to the age group 6 and the binary code of his/her symptoms is 10011111. Therefore, the patient digital code, symptom label and patient label are 6-10011111, 249 and 1524, respectively. The application calculates the probability factors in two ways, directly from the database and using the prediction model. The database shows that the patient has a 20% chance to be admitted to ICU, a 20% chance to be admitted to hospital but not to ICU, a 60% chance for non hospitalization, a 100% chance of recovery with a recovery period of 2.353 weeks. The prediction model shows that the patient has a 19.99% (8.679% to 42.23%) chance to be admitted to ICU, a 23.95% (8.244% to 41.29%) chance to be admitted to hospital but not to ICU, a 56.07% (16.48% to 83.08%) chance for non hospitalization, a 96.56% (82.16% to 100%) chance of recovery with recovery period 2.599 (1.734 to 3.793) weeks.

**Figure 5.**
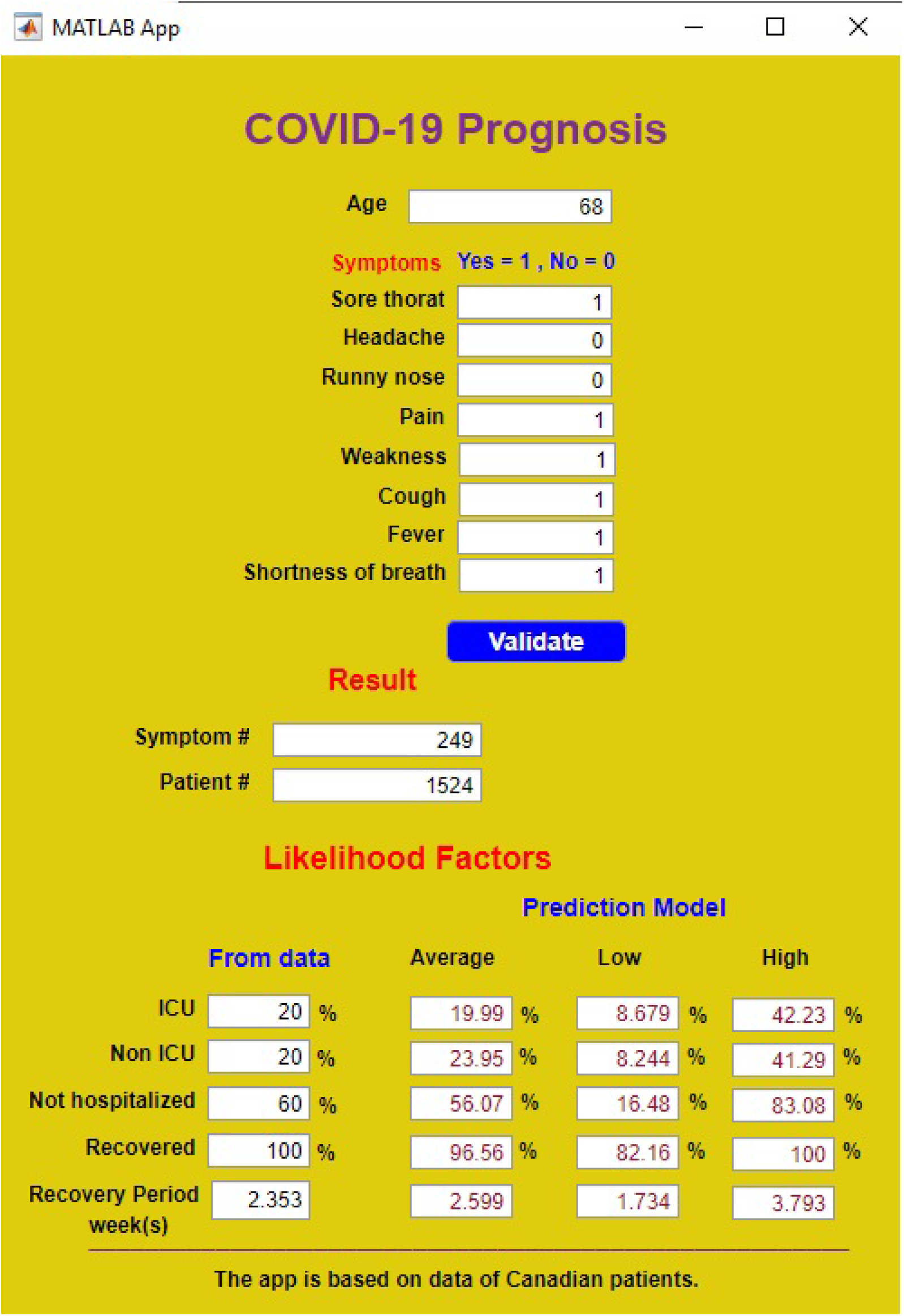
Sketch of the proposed app.

## Discussion

In this paper, we extract the useful information from the database, merely on the basis of age and eight different symptoms, which are represented by the scattered diagrams (Figs. 1 to 4). Some patients in the database have pre-existing illness such as respiratory disease, diabetes, etc. (the database does not provide that type of information) which may be the possible reason of the undulatory behaviour of those diagrams. Suppose two patient groups, patient labels 1524 (patient digital code 6-10011111) and 1525 (patient digital code 6-01011111), and patient group 1524 has more chronic patients than patient group 1525. If this happen, as a consequence, the percentage of ICU and hospitalized patients in the patient group 1524 will be higher than that of the patient group 1525. To overcome the wavy nature of the outcomes, we introduce a prediction model and obtain estimated values with possible ranges.

The application will provide a rapid digital prognosis and at no cost, based on the available database and the prediction model, for COVID-19 patients. This will be useful for health professionals, patients and their families. People related to the COVID-19 patient will be informed and prepared for clinical requirements. People can check the likelihood factors of the patients at each stage, on and off of any symptoms, and take the necessary actions. Using the same methodology and required database, we can develop a similar application for any other infectious disease.

## Limitation and Improvement

The lack of information is the only limitation of the present study, and there are some scopes to improve the outcomes computed by the application using a database that contains more specific information. In this paper, the prediction based on just eight symptoms and age is due to lack of data. However, for a finer prediction, we need to consider more symptoms as well as gender, so we simply need a database that can provide a sufficient amount of such information. For example, if we consider 255 variations of symptoms, eight different age groups and gender, we need a database containing sufficient number of patients information for each group of 255 *×* 8 *×*2 patients groups.

Here, we consider symptoms either present or absent. However, for a more accurate prediction, we need to know the types of the symptoms. For example, instead of fever present or absent if we obtain a database with a sufficiently large size that provides the information of the stages of a fever, such as intermittent, remittent, continuous or sustained, hectic, relapsing and no fever, could generate more groups of patients. If the symptoms have five or six variations instead of just two, used in the present study, we can consider pentagonal or hexagonal system instead of binary system.

Suppose that the digital codes of two patients are the same, i.e., the same age groups and symptoms, but one of them has diabetes. Therefore, these two cases are distinct. However, in the present study there is no such scope to consider the underline health status of the patient, as the database does not provide the patient’s medical history. Including patient underline medical conditions as a variable, such as symptom label, could improve the prognosis.

## Methods

In this section, we introduce a simple algorithm to classify the patients, using an integer code, according to their symptoms and age. The main concept is to assign an integer number correspond to multiple symptoms. Due to lack of enough data of COVID-19 patients in Canada, the present study is not gender-based, and it is confined to eight symptoms.

### Data collection

In this work, we use a preliminary dataset^41^, provided by Statistics Canada (StatCan) in collaboration with the Public Health Agency of Canada (PHAC), as a source of information for the proposed COVID-19 application. The data published by StatCan contains cases for which the provincial or territorial public health authority submitted detailed case information to PHAC. The federal, provincial and territorial governments of Canada have agreed on a Common Case Report Form (CRF) that will be used to report cases to PHAC. The available datasets are preliminary and subject to changes as the information is regularly updated by the provinces and territories. The app could adopt another database with similar information as a source file. Unfortunately, information on symptoms was removed from the dataset as of March 2021, as the information was no longer collected in the CRF because the historical information was incomplete. Hence, we will use the database as of February 2021, cases of 666,854 people with their age group, symptoms, etc. However, the case histories of only 156,988 patients are useful for the present study; other recorded cases are incomplete.

### Symptom label

In the current context, we will consider eight symptoms: sore throat (St), headache (H), runny nose (Rn), pain (P), weakness (W), cough (C), fever (F) and shortness of breath (Sb). An eight digit binary number *b*_8_*b*_7_*b*_6_*b*_5_*b*_4_*b*_3_*b*_2_*b*_1_, referred as a *code*, can be associated to present those eight symptoms such that *b*_1_ = 1 in the presence of the symptom sore throat and *b*_1_ = 0 otherwise; *b*_2_, …, *b*_8_ play the same role as *b*_1_ for other symptoms. The eight digit binary number corresponds to a decimal number (integer) *σ*, defined as a symptom label (Fig. 6(a)), which can be expressed as

**Figure 6.**
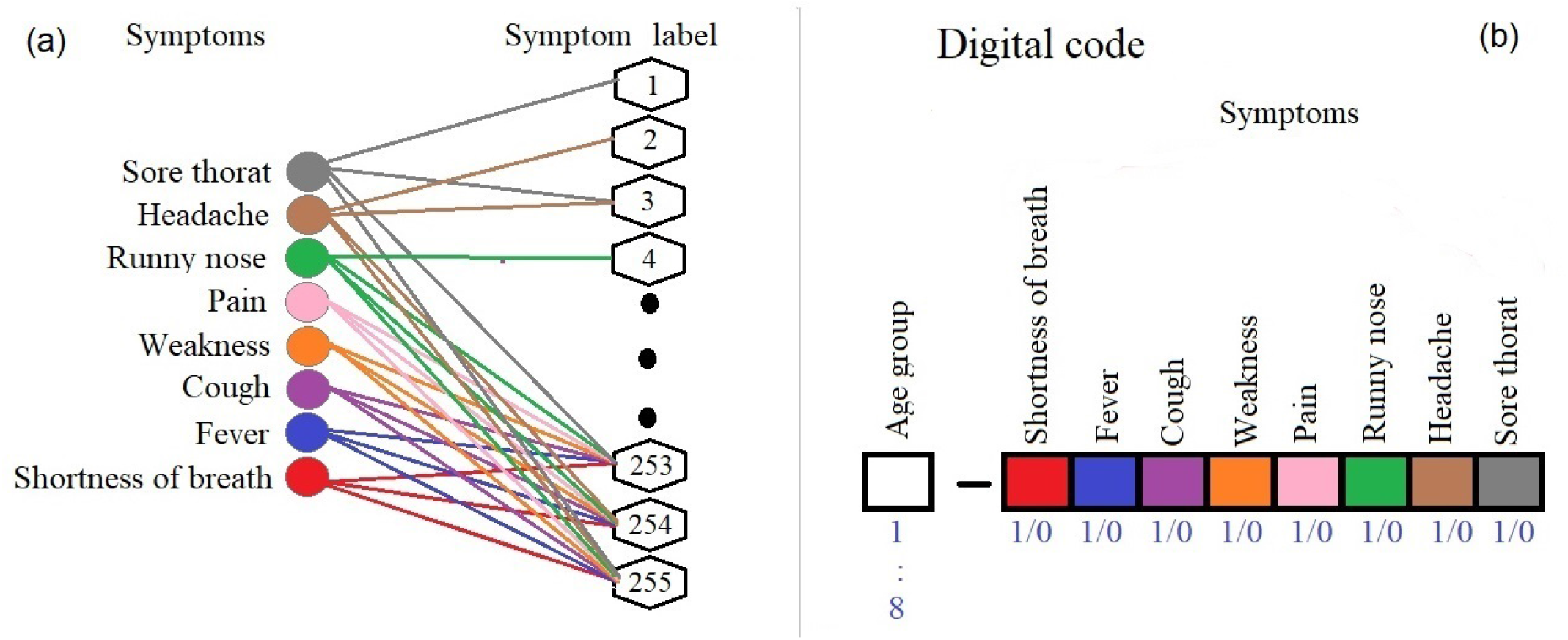
Schematic diagram for symptom label, patient label and patient digital code.

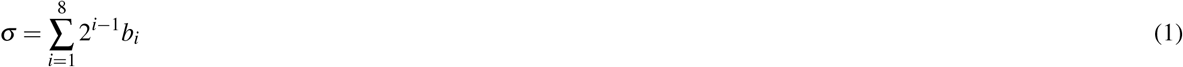

where 1 *≤ σ ≤* 2^8^ *-* 1.

Table 1 shows that an integer number, defined as symptom label, indicates the occurrence of multiple symptoms. For example, symptom label 1 corresponds to the only symptom sore throat; if a patient has all those eight symptoms, his / her symptom label is 255. Here, we assign symptom label such that low symptom labels correspond to the less severe cases, and high symptom labels indicate more severe cases.

**Table 1.** Symptom label corresponding eight different symptoms. Y (N) represents the presence (absence) of a symptom.

| Symptoms |  |  |  |  |  |  |  | Binary number | Symptom label |
| --- | --- | --- | --- | --- | --- | --- | --- | --- | --- |
| Sb | F | C | W | P | Rn | H | St |  |  |
| N | N | N | N | N | N | N | Y | 00000001 | 1 |
| N | N | Y | Y | N | N | N | Y | 00110001 | 49 |
| Y | N | Y | N | N | Y | Y | N | 10100110 | 166 |
| Y | Y | Y | Y | Y | Y | Y | Y | 11111111 | 255 |

### Patient label

Including age-groups, we can obtain patient labels *ρ*, defined as

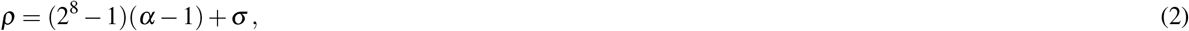

where *α* = 1, 2,*…*, 8 for eight different age-groups.

We segregate all the patients into 2^8^ *-*1 = 255 groups according to the symptom labels and into 8 *×*255 = 2040 groups according to the patient labels. For example, if one patient belongs to the age group 6 with symptom label 49, his / her patient label is 6*×* 255 + 49 = 1324. The maximum patient label, 2040, represents all the possible cases based on symptoms and age. We can also define patient digital code (Fig. 6(b)) according to patient age group and symptoms. As an illustration, suppose digital code of a patient is 4 *-*10100110 which indicates that the patient belongs to age group 4 with symptom label 166 (Table 1). The digital code of a patient can use as input data for the digital prognosis application.

We develop a matlab code to extract patient likelihood factors such as the percentage of patients being admitted to ICU, the percentage of patients being admitted to hospital but not to the ICU, etc. from the available database. Finally, we generate an output file of those 2040 cases that is used in the COVID-19 app.

### Prediction model

We assign the symptom label such that less (more) severe condition represents by the low (high) symptom label (Tabel 1). Generally, when we extract some information from the database, for example the number of hospitalized patients as a function of symptom label, we observe a scatter diagram. In this situation, to obtain consistent outcomes, we need a prediction model. We consider a prediction model *P* as a power function of symptom label *σ*

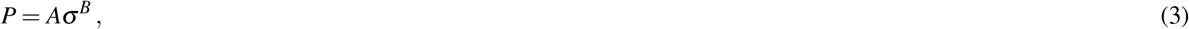

where the real parameters A and B, which are functions of age groups, are estinated thanks to a least square method. The prediction model hence provides an overall average.

### Range of the prediction

To calculate the range of the prediction model, we first decompose the domain of the symptom labels *S* into several subdomains.

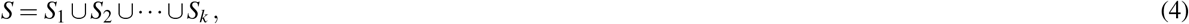

where *S* = {1, 2, … 255} and *S* _*j*_ for *j* = 1, 2, …, *k* are *k* subdomains of *S*. Now we find the minimum and maximum values of a physical observable, for example number of hospitalized patients, lies in each subdomain *S* _*j*_ for *j* = 1, 2, …, *k*. Fitting those minimum values of a physical observable with the curve *P*_*min*_, defined as

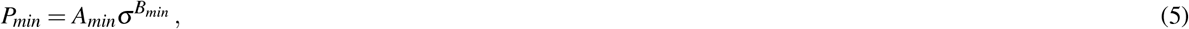

where the real parameters *A*_*min*_ and *B*_*min*_ are functions of the age groups we get the lower range of the prediction. Similarly using the maximum values of a physical observable and the curve *P*_*max*_, defined as

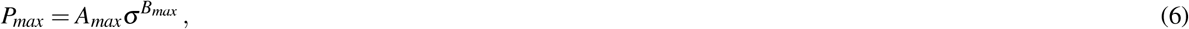

where the real parameters *A*_*max*_ and *B*_*max*_ are functions of the age groups, we get the upper range of the prediction.

### Development of the app

In our present study total number of patient labels is 2040 i.e., we have a total 2040 distinct patient groups according to their symptoms and age groups. We generate some data files of 2040 groups of patients such as the probability of patients being admitted to the ICU, the average recovery period, etc. We also gather those informations from the prediction models *P, P*_*min*_ and *P*_*max*_. We develop an application using all those data files. Each of the 2040 distinct groups indicates a specific age group and symptom pattern. Thus if someone provides the information about his/her age group and symptoms, the application will provide some outcome probabilities.

## Data Availability

https://www150.statcan.gc.ca/n1/en/catalogue/13260003

## Data availability

The data used to estimate the model parameters are publicly available and are available with the code.

## Code availability

All code is available in the GitHub repository for the project at https://github.com/SPAUL2021/COVID-19_Prognosis

## Acknowledgements

This research is supported by the Natural Sciences and Engineering Research Council of Canada (NSERC).

## Author contributions statement

S.P. has derived the prediction model, has developed the matlab code and application, has analyzed the calculated results, and has prepared all figures and table. S.P. and E.L. have drafted the original article. Both authors have contributed to the editing of the article. Both authors have read and approved the final article.

## Competing interests

The authors declare that they have no competing interests.

## Additional information

Correspondence and requests for materials should be addressed to S.P.

## Notes

### Competing Interest Statement

The authors have declared no competing interest.

### Author Declarations

Data of Canadian COVID-19 patients are publicly available in the webpage of statistical Canada.

